# Serum anti-nucleocapsid antibody level induced after primary infection is an immunological surrogate of protection against SARS-CoV-2 re-infection in hybrid immunity holders

**DOI:** 10.1101/2024.06.05.24308479

**Authors:** Sho Miyamoto, Koki Numakura, Ryo Kinoshita, Takeshi Arashiro, Hiromizu Takahashi, Hiromi Hibino, Minako Hayakawa, Takayuki Kanno, Akiko Sataka, Akira Ainai, Satoru Arai, Motoi Suzuki, Daisuke Yoneoka, Takaji Wakita, Tadaki Suzuki

## Abstract

**Background:** In 2024, there was quite high seroprevalence of anti-spike (S) protein antibodies against SARS-CoV-2 in Japanese adults, owing to the high vaccination coverage by spike-based vaccines. Nevertheless, the COVID-19 epidemic continues, albeit with low rates of severe illness, and hybrid immunity holders are becoming more common in these populations. It is necessary to determine the immunological protection correlates against SARS-CoV-2 re-infection in individuals with hybrid immunity because the currently available immune correlates were established by analyzing individuals possessing vaccine-induced immunity only.

**Methods:** We conducted an ad hoc prospective cohort study to measure serum anti-SARS-CoV-2 antibody levels in 4,496 Japanese adults as part of the national COVID-19 seroepidemiological survey. This ad hoc study evaluated the correlation between anti-S and anti-nucleocapsid (N) antibody levels at the first visit and their effectiveness in infection prevention until the second visit, including undiagnosed re-infections during the Omicron BA.5 epidemic period from December 2022 to March 2023.

**Findings:** We assessed the combined effect of anti-N and anti-S antibody levels and found that the reduced infection risk associated with anti-S antibody levels was limited. Contrastingly, higher levels of anti-N antibodies were strongly linked to a reduced infection risk in the entire cohort and in individuals with hybrid immunity.

**Interpretation:** We demonstrate a high correlation between reduced re-infection risk in hybrid immunity holders and high serum anti-N antibody levels, highlighting its potential as an immunological surrogate of protection against SARS-CoV-2 re-infection. The findings indicate that individuals with hybrid immunity are protected by a distinct form of immunity, beyond the presence of serum anti-S antibodies, which correlates with serum anti-N antibody levels.

**Funding:** The national COVID-19 seroepidemiological survey as a public health investigation was funded by the Ministry of Health, Labour and Welfare of Japan (MHLW). The ad hoc study based on the survey data as a research activity was funded by the Japan Agency for Medical Research and Development (AMED).

**Research in context:** *Evidence before this study:* We searched PubMed for studies published between January 1, 2022, and April 18, 2024, using the search terms “SARS-CoV-2” in combination with the search terms “antibody,” “Omicron,” AND “Correlate(s) of Protection,” with no language restrictions. Studies on the correlates of protection (CoP) using antibody titers to prevent Omicron infection have primarily been performed during Omicron BA.1/2 waves. One report indicated serum correlates of protection involving anti-spike (S) antibodies against Omicron BA.5, but the anti-S antibody titer thresholds varied according to previous infection histories. The investigation of quantitative immunological markers that serve as correlates of protection against infection among populations with various immune histories through vaccination and infection should include asymptomatic or undiagnosed re-infected cases, which would be useful for the development of next-generation COVID-19 vaccines that would control future COVID-19 epidemics. However, the immune correlates of protection against re-infection, especially among hybrid immunity holders with a history of infections and vaccination, remains unclear.

*Added value of this study:* Our study evaluated immunological markers for infection prevention in adults with both vaccination and infection histories during the Omicron sublineage epidemic period. The reduction in re-infection risk during the Omicron BA.5 epidemic period correlated with higher anti-nucleocapsid (N) antibody levels. Conversely, anti-S antibody titers induced by both vaccines and infections were less strongly correlated with protection. These results may account for the variation in anti-S antibody titers’ effectiveness in protecting against Omicron sublineages, highlighting the usefulness of anti-N antibody levels for estimating the antiviral immunity level in hybrid immunity holders, the majority of the population with high vaccination coverage.

*Implications of all the available evidence:* Previously established immunological correlates for the prevention of SARS-CoV-2 infection are serum anti-S antibody levels and neutralization titers induced by vaccination or infection. In contrast, serum anti-N antibody responses are considered to be immune responses induced by infection. Our findings suggest that infection-induced anti-N antibody levels represent a non-mechanical immunological surrogate for protection against re-infection. According to the study’s results, people with hybrid immunity have an unique immunity that correlates with serum anti-N antibody levels above and beyond the presence of serum anti-S antibodies, suggesting the potential for the development of a next-generation COVID-19 vaccine that can induce more effective immunity by mimicking hybrid immunity.

## Introduction

Several spike-based vaccines against SARS-CoV-2 with mRNA or viral vector modalities were developed during the early COVID-19 pandemic and showed high efficacy in early clinical trials and during the pre-Omicron epidemic period. However, since the emergence of Omicron variants at the end of 2021, there have been continuous reports of Omicron sublineages with high resistance to humoral immunity induced by spike-based vaccines,^1,2^ leading to a decline in vaccine effectiveness against infection.^3,4^ Anti-spike (S) antibody titers against the ancestral strain induced by vaccination were identified as immunological correlates for protection against SARS-CoV-2 infection and severe disease during the pre-Omicron epidemic period.^5–7^ While the prevention of severe disease through vaccination has been confirmed even during the Omicron BA.1/2 and BA.4/5 epidemic periods ^8^, higher anti-spike (S) antibody titers against the ancestral strain were required for protection against infection during these periods compared to the pre-Omicron epidemic period.^9–12^ Omicron sublineages tend to be selected for mutations with high humoral immune evasion capabilities,^13^ and the protective effect of anti-S antibody titers against the ancestral strain needs to be re-evaluated in response to changes in the antigenicity of the emerging variants.^14^ Recently, with the increase in the proportion of infected individuals, it has been reported that not only the neutralizing antibody titers and anti-S antibody titers induced by vaccination but also past infection history are significantly associated with a reduction in infection risk.^15,16^ Additionally, recent reports have shown that mucosal secretory IgA antibody levels, elicited after infection, are associated with the prevention of SARS-CoV-2 infection and shedding.^17,18^ Therefore, to accurately estimate the extent of the COVID-19 epidemic in the post-COVID-19 pandemic era, it is necessary to determine the immunological correlates of protection against SARS-CoV-2 re-infection in individuals with hybrid immunity due to both vaccination and infection.^19^

Evaluating the accurate potential of immunological correlates for preventing infection requires the assessment of asymptomatic and undiagnosed re-infected cases, which also influence the COVID-19 epidemic dynamics. Recently, undiagnosed re-infections were reported to be prevalent, implying that analyses focusing only on symptomatic infections risk underestimating the re-infection risk.^20,21^ Against this backdrop, this study conducted a nationwide cohort survey in Japan that involved two blood samplings and antibody tests during the Omicron BA.5 epidemic period (https://cov-spectrum.org) from December 2022 to March 2023. By identifying infected individuals through diagnosis and seroconversion of infection-derived anti-nucleocapsid (N) antibodies, we included undiagnosed primary infections and re-infections as newly infected cases during the study period. We then evaluated the infection prevention efficacy of anti-S antibody titers and the combined effect of infection-related anti-N antibody levels during the Omicron BA.5 epidemic period by analyzing the association of combined serum anti-S and anti-N antibody levels with newly infected cases.

## Results

### Characteristics of the study participants and the antibody responses in newly infected cases

Of the 15,000 invitees for the national COVID-19 seroepidemiological survey conducted as a public health investigation, 8,157 participated in the December 2022 survey and 5,627 participated in the February to March 2023 survey (response rate: 37·5%; **Fig. 1**). We targeted the 5,627 individuals who participated in both surveys. Those who received any COVID-19 vaccine during the observation period were excluded (1,044 individuals) to eliminate the influence of the vaccination immune response during the study period. In addition, 87 individuals with a history of infection within 30 days before the initial test date were excluded (87 individuals) because the antibody response up to 30 days after infection is dynamic and difficult to accurately assess at this blood collection interval. Therefore, 4,496 individuals were enrolled for the ad hoc study. Additionally, hybrid immunity holders were defined as vaccinated individuals with a history of prior diagnoses of COVID-19 or the anti-N antibodies positive (anti-N antibody levels > 1·0; cutoff index, [COI]) at the initial test.

**Fig. 1.**
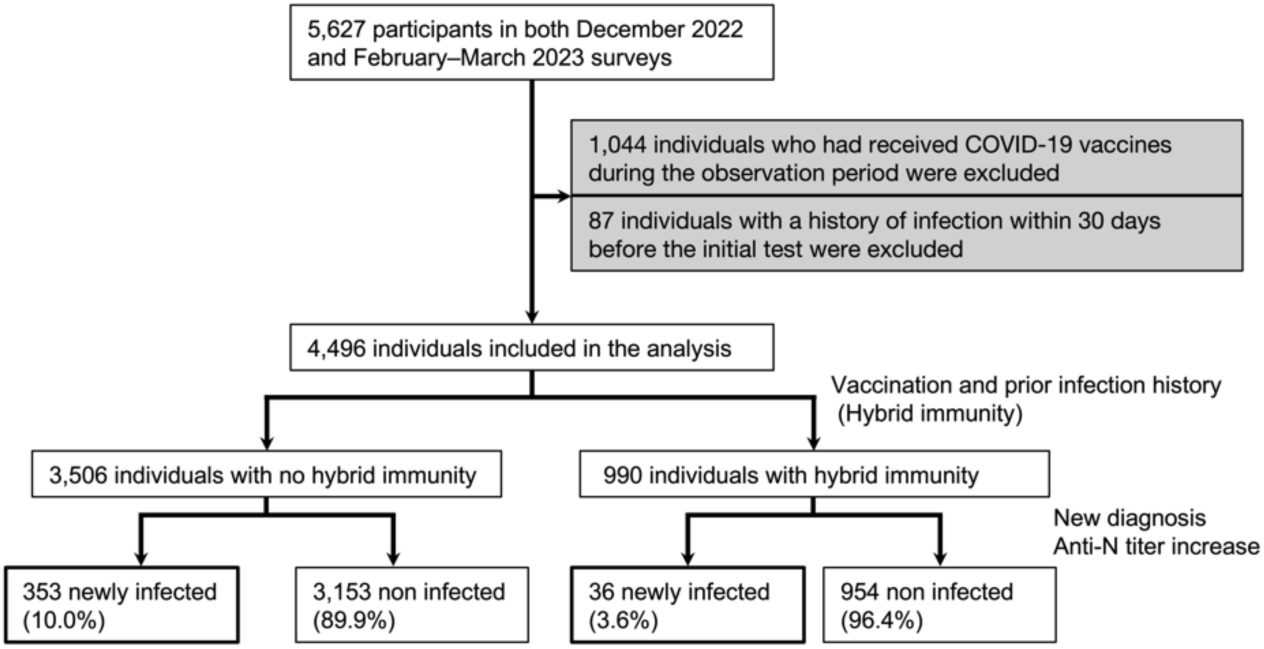
Flow diagram of the study design and the included participants.

Among participants without a history of infection at the time of the initial test, those who were diagnosed with COVID-19 after the initial test date or whose anti-N antibodies were seroconverted were regarded as newly infected cases. The median fold increase in anti-N antibody levels from the initial to the second test in newly infected cases with no hybrid immunity at the initial test date was 288·3 (IQR, 150·0–784·0; **Fig. 2A**). In contrast, anti-S antibody titers in newly infected cases with no hybrid immunity at the initial test showed a limited increase (5·6-fold increase; IQR, 2·3–10·9; **Fig. 2B**) because the baseline level of anti-S antibody titers in this cohort was comparable with that of the booster vaccination due to high booster vaccination rate (71.2%; **Table 1**). Additionally, the correlation between the anti-S and anti-N antibody levels among all participants was low (**Fig. 2C**). This suggests that the anti-N antibody response induced by infection during the Omicron BA.5 endemic period is significantly larger and superior for the detection of infection compared to the anti-S antibody response induced by infection in this cohort.

**Fig. 2.**
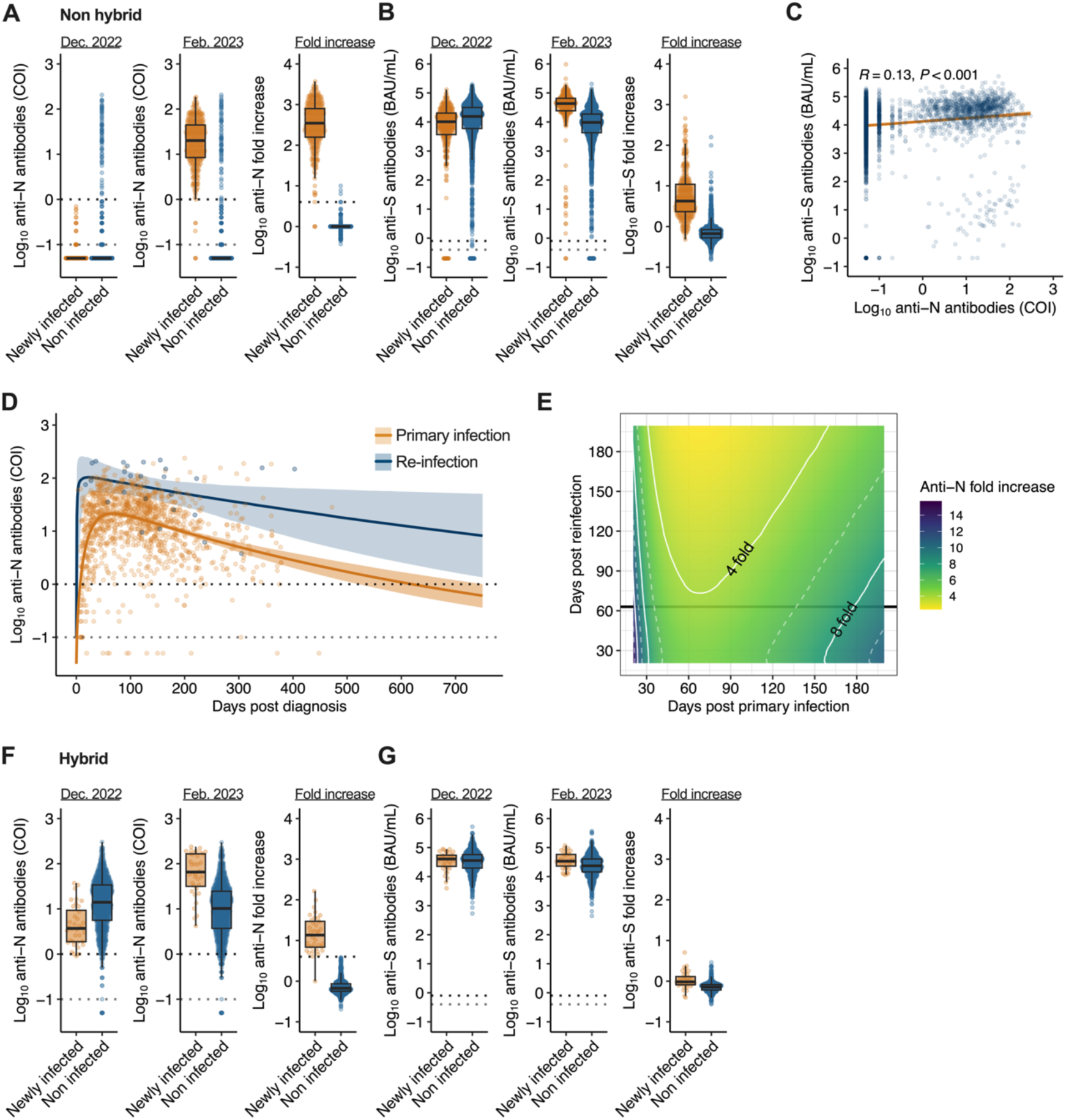
Serum anti-S and anti-N antibody levels in the presence or absence of new infections and the anti-N antibody level dynamics for each infection history. (A, B) The antibody levels and their fold increase for participants without hybrid immunity at baseline (Dec. 2022) and the end of observation (Feb. 2023). (A) Anti-N antibody levels and (B) anti-S antibody titers. (C) Correlation between serum anti-S and anti-N antibody levels for all participants. Correlation coefficients and P values, along with regression lines and 95% confidence intervals, are shown. (D) A model of the dynamics of anti-N antibody levels in primary infection and re-infection. Each data point, along with the median and 95% credible intervals (ribbon) for the mean anti-N antibody levels, are shown. (E) Estimation of the fold increase in anti-N antibody levels induced by re-infection. The fold increase was calculated from the ratio of estimated anti-N antibody levels on the days post primary diagnosis (infection) at re-infection to the titers at any days post-re-infection. The white dotted and solid lines show a 2x and 4x fold increase, respectively, in the anti-N antibody level. The black line shows the median observation period (63 days). (F, G) The baseline (Dec. 2022) and end-of-observation (Feb. 2023) antibody titers and their fold increase in individuals with hybrid immunity. (F) Anti-N antibody levels and (G) anti-S antibody titers. In antibody levels, the dark gray dotted line indicates the cutoff value according to the manufacturer’s manual, and the light gray dotted line represents the detection limit.

**Table 1.**
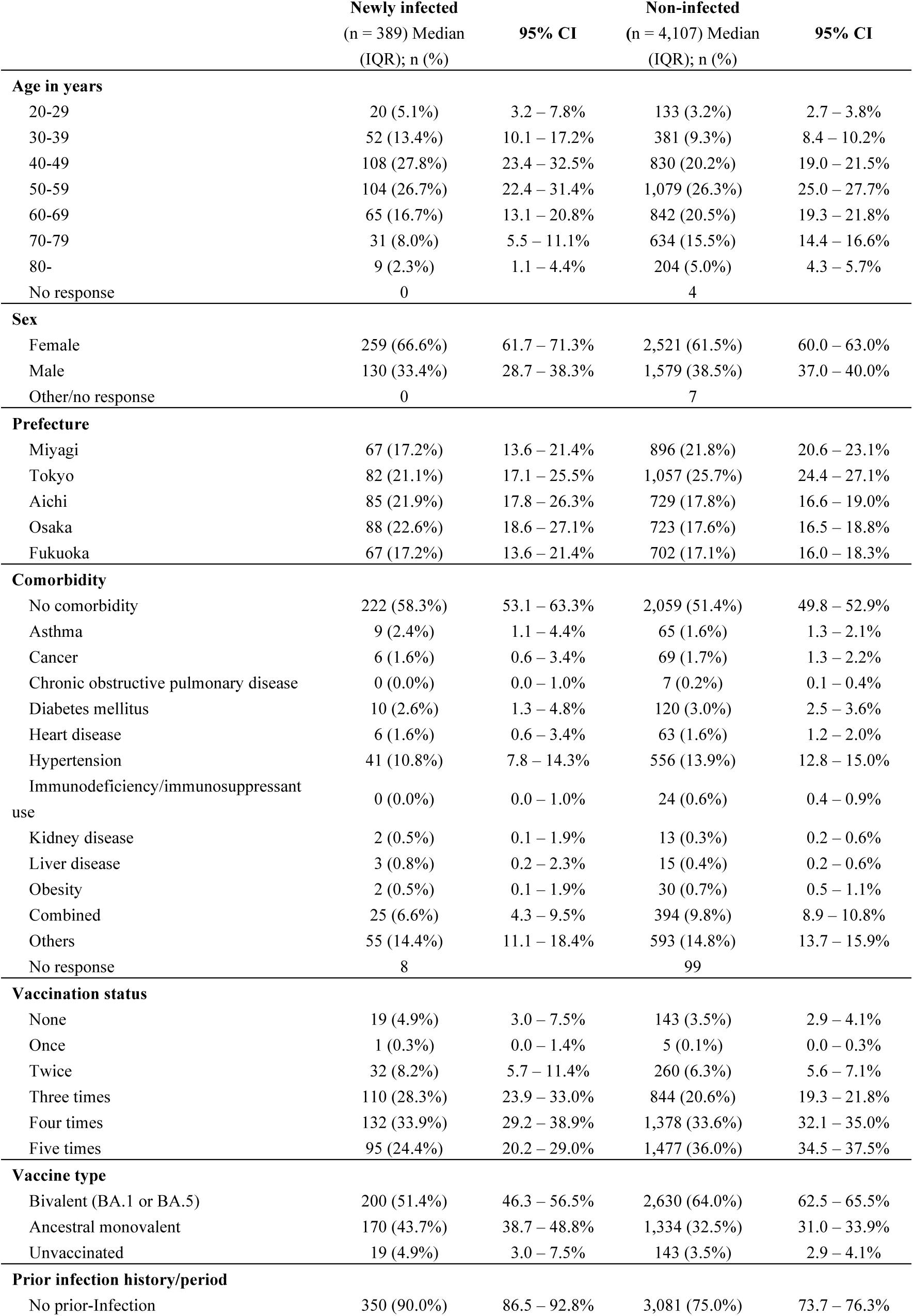
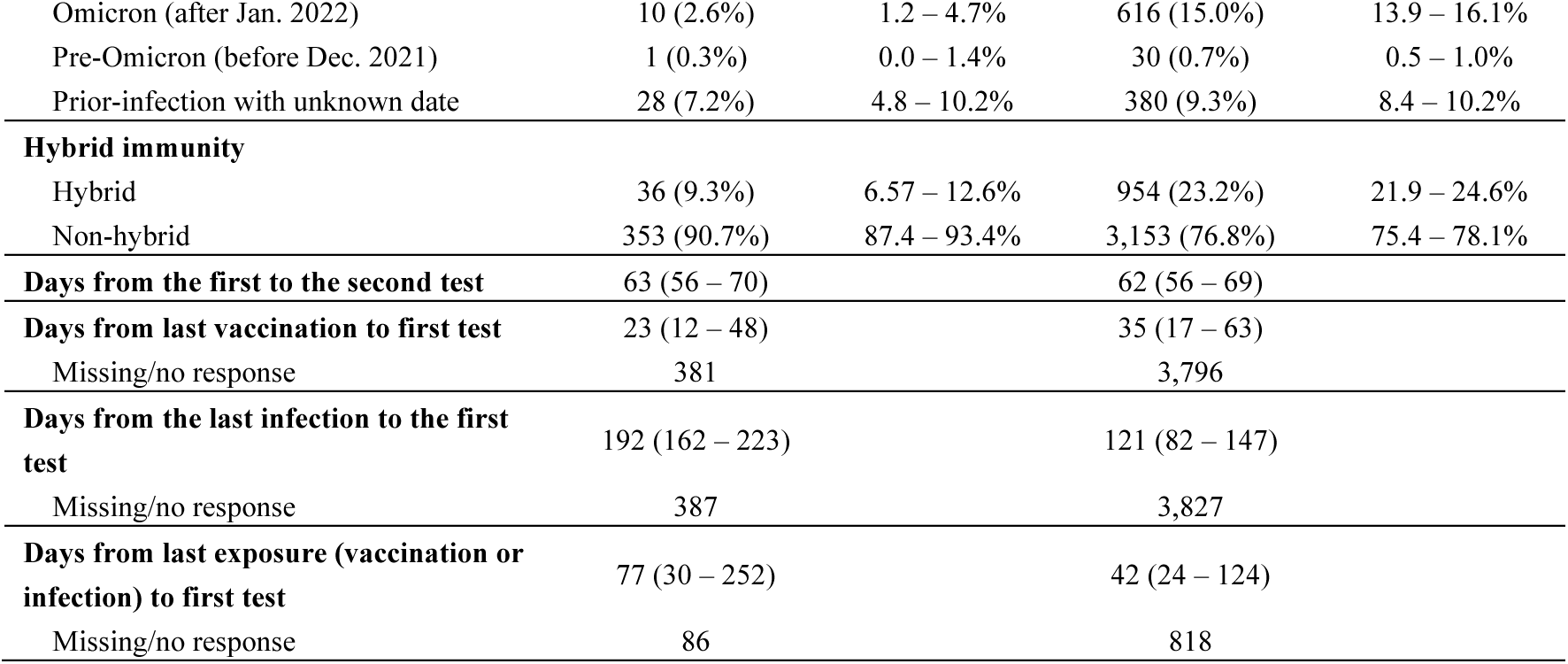
Characteristics of individuals with and without infection during the study period.

|  | Newly infected<br>(n = 389) Median<br>(IQR); n (%) | 95% CI | Non-infected<br>(n = 4,107) Median<br>(IQR); n (%) | 95% CI |
| --- | --- | --- | --- | --- |
| **Age in years** |  |  |  |  |
| 20-29 | 20 (5.1%) | 3.2 – 7.8% | 133 (3.2%) | 2.7 – 3.8% |
| 30-39 | 52 (13.4%) | 10.1 – 17.2% | 381 (9.3%) | 8.4 – 10.2% |
| 40-49 | 108 (27.8%) | 23.4 – 32.5% | 830 (20.2%) | 19.0 – 21.5% |
| 50-59 | 104 (26.7%) | 22.4 – 31.4% | 1,079 (26.3%) | 25.0 – 27.7% |
| 60-69 | 65 (16.7%) | 13.1 – 20.8% | 842 (20.5%) | 19.3 – 21.8% |
| 70-79 | 31 (8.0%) | 5.5 – 11.1% | 634 (15.5%) | 14.4 – 16.6% |
| 80- | 9 (2.3%) | 1.1 – 4.4% | 204 (5.0%) | 4.3 – 5.7% |
| No response | 0 |  | 4 |  |
| **Sex** |  |  |  |  |
| Female | 259 (66.6%) | 61.7 – 71.3% | 2,521 (61.5%) | 60.0 – 63.0% |
| Male | 130 (33.4%) | 28.7 – 38.3% | 1,579 (38.5%) | 37.0 – 40.0% |
| Other/no response | 0 |  | 7 |  |
| **Prefecture** |  |  |  |  |
| Miyagi | 67 (17.2%) | 13.6 – 21.4% | 896 (21.8%) | 20.6 – 23.1% |
| Tokyo | 82 (21.1%) | 17.1 – 25.5% | 1,057 (25.7%) | 24.4 – 27.1% |
| Aichi | 85 (21.9%) | 17.8 – 26.3% | 729 (17.8%) | 16.6 – 19.0% |
| Osaka | 88 (22.6%) | 18.6 – 27.1% | 723 (17.6%) | 16.5 – 18.8% |
| Fukuoka | 67 (17.2%) | 13.6 – 21.4% | 702 (17.1%) | 16.0 – 18.3% |
| **Comorbidity** |  |  |  |  |
| No comorbidity | 222 (58.3%) | 53.1 – 63.3% | 2,059 (51.4%) | 49.8 – 52.9% |
| Asthma | 9 (2.4%) | 1.1 – 4.4% | 65 (1.6%) | 1.3 – 2.1% |
| Cancer | 6 (1.6%) | 0.6 – 3.4% | 69 (1.7%) | 1.3 – 2.2% |
| Chronic obstructive pulmonary disease | 0 (0.0%) | 0.0 – 1.0% | 7 (0.2%) | 0.1 – 0.4% |
| Diabetes mellitus | 10 (2.6%) | 1.3 – 4.8% | 120 (3.0%) | 2.5 – 3.6% |
| Heart disease | 6 (1.6%) | 0.6 – 3.4% | 63 (1.6%) | 1.2 – 2.0% |
| Hypertension | 41 (10.8%) | 7.8 – 14.3% | 556 (13.9%) | 12.8 – 15.0% |
| Immunodeficiency/immunosuppressant use | 0 (0.0%) | 0.0 – 1.0% | 24 (0.6%) | 0.4 – 0.9% |
| Kidney disease | 2 (0.5%) | 0.1 – 1.9% | 13 (0.3%) | 0.2 – 0.6% |
| Liver disease | 3 (0.8%) | 0.2 – 2.3% | 15 (0.4%) | 0.2 – 0.6% |
| Obesity | 2 (0.5%) | 0.1 – 1.9% | 30 (0.7%) | 0.5 – 1.1% |
| Combined | 25 (6.6%) | 4.3 – 9.5% | 394 (9.8%) | 8.9 – 10.8% |
| Others | 55 (14.4%) | 11.1 – 18.4% | 593 (14.8%) | 13.7 – 15.9% |
| No response | 8 |  | 99 |  |
| **Vaccination status** |  |  |  |  |
| None | 19 (4.9%) | 3.0 – 7.5% | 143 (3.5%) | 2.9 – 4.1% |
| Once | 1 (0.3%) | 0.0 – 1.4% | 5 (0.1%) | 0.0 – 0.3% |
| Twice | 32 (8.2%) | 5.7 – 11.4% | 260 (6.3%) | 5.6 – 7.1% |
| Three times | 110 (28.3%) | 23.9 – 33.0% | 844 (20.6%) | 19.3 – 21.8% |
| Four times | 132 (33.9%) | 29.2 – 38.9% | 1,378 (33.6%) | 32.1 – 35.0% |
| Five times | 95 (24.4%) | 20.2 – 29.0% | 1,477 (36.0%) | 34.5 – 37.5% |
| **Vaccine type** |  |  |  |  |
| Bivalent (BA.1 or BA.5) | 200 (51.4%) | 46.3 – 56.5% | 2,630 (64.0%) | 62.5 – 65.5% |
| Ancestral monovalent | 170 (43.7%) | 38.7 – 48.8% | 1,334 (32.5%) | 31.0 – 33.9% |
| Unvaccinated | 19 (4.9%) | 3.0 – 7.5% | 143 (3.5%) | 2.9 – 4.1% |
| **Prior infection history/period** |  |  |  |  |
| No prior-Infection | 350 (90.0%) | 86.5 – 92.8% | 3,081 (75.0%) | 73.7 – 76.3% |
| Omicron (after Jan. 2022) | 10 (2.6%) | 1.2 – 4.7% | 616 (15.0%) | 13.9 – 16.1% |
| Pre-Omicron (before Dec. 2021) | 1 (0.3%) | 0.0 – 1.4% | 30 (0.7%) | 0.5 – 1.0% |
| Prior-infection with unknown date | 28 (7.2%) | 4.8 – 10.2% | 380 (9.3%) | 8.4 – 10.2% |
| **Hybrid immunity** |  |  |  |  |
| Hybrid | 36 (9.3%) | 6.57 – 12.6% | 954 (23.2%) | 21.9 – 24.6% |
| Non-hybrid | 353 (90.7%) | 87.4 – 93.4% | 3,153 (76.8%) | 75.4 – 78.1% |
| **Days from the first to the second test** | 63 (56 – 70) |  | 62 (56 – 69) |  |
| **Days from last vaccination to first test** | 23 (12 – 48) |  | 35 (17 – 63) |  |
| Missing/no response | 381 |  | 3,796 |  |
| **Days from the last infection to the first test** | 192 (162 – 223) |  | 121 (82 – 147) |  |
| Missing/no response | 387 |  | 3,827 |  |
| **Days from last exposure (vaccination or infection) to first test** | 77 (30 – 252) |  | 42 (24 – 124) |  |
| Missing/no response | 86 |  | 818 |  |

Next, we estimated the dynamics of anti-N antibody responses in individuals diagnosed with primary infection or re-infection to understand the differences in anti-N antibody responses. We applied a statistical model for the anti-N antibody response dynamics after diagnosis for all primary infected and re-infected cases using data obtained from cases with only the listed diagnosis date (primary infection, n = 1218; re-infection, n = 26) (**Fig. 2D**). For primary infected cases, anti-N titers were estimated to peak 69 days post-diagnosis and drop below the positive threshold of 1·0 at 621 days (95% credible interval (CredI), 522–754). For re-infected cases, the peak level of anti-N-antibody was estimated as 4·8 times higher than that of primary infected cases, and the duration of antibody levels above 1·0 COI was estimated to be longer than that of primary infected cases. Using this model, we calculated the fold-increase in anti-N antibody levels from the pre-reinfection level to the level up to 200 days post-re-infection (**Fig. 2E**). Because the interval from the initial to the second test in this cohort was approximately 2 months the lowest fold-increase in anti-N antibody level from the pre-reinfection state to 60 days after re-infection was estimated to be four-fold or higher. These anti-N antibody response dynamics in those who were re-infected were consistent with previous reports using the same anti-N antibody detection kit.^22,23^ These results suggest that a four-fold increase in anti-N antibody levels from the initial test to the second test in this cohort was considered a re-infection during the study period. In individuals with hybrid immunity, the anti-N antibody levels of newly infected cases were lower in the initial test but higher in the second test than those in non-infected cases (**Fig. 2F**). The median anti-N antibody levels’ fold-increase from the initial to second tests in cases with COVID-19 diagnosis during the study period was 14·0 (IQR, 6·8–29·6) for those with hybrid immunity before the study (**Fig. 2F**). In contrast, among newly-infected cases with hybrid immunity, no clear increase in anti-S antibody titers from the initial to the second tests was observed, and anti-S antibody titers in cases with COVID-19 diagnosis during the study period hardly changed from the initial to second tests (1·1-fold increase; IQR, 0·8–1·3; **Fig. 2G**). Taken together, these results justify the use of the criteria of a four-fold increase in anti-N antibody levels during the study period to identify newly infected cases with or without hybrid immunity before the study period, including undiagnosed cases.

The characteristics of newly infected and non-infected cases are presented in Table 1, and the characteristics of newly infected cases acquiring hybrid immunity before the study period are shown in Table 2. Among the newly infected cases, the proportions of elderly individuals aged over 70 years, those with comorbidities, individuals who had received five vaccine doses, those who had received the Omicron-adapted bivalent vaccine, and individuals with a history of infection were relatively low (**Table 1**). No apparent differences were observed between patients with or without hybrid immunity regarding comorbidity or vaccination status (**Table 2**). Of the patients without hybrid immunity, 51.0% were diagnosed with COVID-19 during the study period. In contrast, only 5.6% of the re-infected patients with hybrid immunity were diagnosed with COVID-19, indicating that most of the re-infected patients with hybrid immunity were undiagnosed and undetectable without serological testing (**Table 2**). Similarly, only 5.6% of the re-infected cases with hybrid immunity were symptomatic, suggesting that most re-infected cases were asymptomatic and less likely to be recognized by symptom-based case identification (**Table 2**).

**Table 2.** Characteristics of newly infected cases during the study period.

|  | Non-hybrid<br>(n = 353) n (%) | 95% CI | Hybrid<br>(n = 36) n (%) | 95% CI |
| --- | --- | --- | --- | --- |
| **Age in years** |  |  |  |  |
| 20-29 | 17 (4.8%) | 2.8 – 7.6% | 3 (8.3%) | 1.8 – 22.5% |
| 30-39 | 47 (13.3%) | 10.0 – 17.3% | 5 (13.9%) | 4.7 – 29.5% |
| 40-49 | 96 (27.2%) | 22.6 – 32.2% | 12 (33.3%) | 18.6 – 51.0% |
| 50-59 | 96 (27.2%) | 22.6 – 32.2% | 8 (22.2%) | 10.1 – 39.2% |
| 60-69 | 59 (16.7%) | 13.0 – 21.0% | 6 (16.7%) | 6.4 – 32.8% |
| 70-79 | 30 (8.5%) | 5.8 – 11.9% | 1 (2.8%) | 0.1 – 14.5% |
| 80- | 8 (2.3%) | 1.0 – 4.4% | 1 (2.8%) | 0.1 – 14.5% |
| **Sex** |  |  |  |  |
| Female | 236 (66.9%) | 61.7 – 71.7% | 23 (63.9%) | 46.2 – 79.2% |
| Male | 117 (33.1%) | 28.3 – 38.3% | 13 (36.1%) | 20.8 – 53.8% |
| **Prefecture** |  |  |  |  |
| Miyagi | 65 (18.4%) | 14.5 – 22.9% | 2 (5.6%) | 0.7 – 18.7% |
| Tokyo | 75 (21.2%) | 17.1 – 25.9% | 7 (19.4%) | 8.2 – 36.0% |
| Aichi | 76 (21.5%) | 17.4 – 26.2% | 9 (25%) | 12.1 – 42.2% |
| Osaka | 77 (21.8%) | 17.6 – 26.5% | 11 (30.6%) | 16.3 – 48.1% |
| Fukuoka | 60 (17%) | 13.2 – 21.3% | 7 (19.4%) | 8.2 – 36.0% |
| **Vaccination status** |  |  |  |  |
| None | 19 (5.4%) | 3.27 – 8.28% | 0 (0.0%) | 0.0 – 9.7% |
| Once | 1 (0.3%) | 0.0 – 1.6% | 0 (0.0%) | 0.0 – 9.7% |
| Twice | 30 (8.5%) | 5.8 – 11.9% | 2 (5.6%) | 0.7 – 18.7% |
| Three times | 97 (27.5%) | 22.9 – 32.5% | 13 (36.1%) | 20.8 – 53.8% |
| Four times | 121 (34.3%) | 29.3 – 39.5% | 11 (30.6%) | 16.3 – 48.1% |
| Five times | 85 (24.1%) | 19.7 – 28.9% | 10 (27.8%) | 14.2 – 45.2% |
| **Comorbidity** |  |  |  |  |
| No comorbidity | 195 (56.5%) | 51.1 – 61.8% | 27 (75%) | 57.8 – 87.9% |
| Asthma | 9 (2.6%) | 1.2 – 4.9% | 0 (0.0%) | 0.0 – 9.7% |
| Cancer | 6 (1.7%) | 0.6 – 3.8% | 0 (0.0%) | 0.0 – 9.7% |
| Chronic obstructive<br>pulmonary disease | 0 (0.0%) | 0.0 – 1.0% | 0 (0.0%) | 0.0 – 9.7% |
| Diabetes mellitus | 9 (2.6%) | 1.2 – 4.9% | 1 (2.8%) | 0.1 – 14.5% |
| Heart disease | 6 (1.7%) | 0.6 – 3.8% | 0 (0.0%) | 0.0 – 9.7% |
| Hypertension | 39 (11.3%) | 8.2 – 15.1% | 2 (5.6%) | 0.7 – 18.7% |
| Immunodeficiency/immunosu<br>ppressant use | 0 (0.0%) | 0.0 – 1.0% | 0 (0.0%) | 0.0 – 9.7% |
| Kidney disease | 2 (0.6%) | 0.1 – 2.1% | 0 (0.0%) | 0.0 – 9.7% |
| Liver disease | 2 (0.6%) | 0.1 – 2.1% | 1 (2.8%) | 0.1 – 14.5% |
| Obesity | 1 (0.3%) | 0.0 – 1.6% | 1 (2.8%) | 0.1 – 14.5% |
| Combined | 25 (7.2%) | 4.7 – 10.5% | 0 (0.0%) | 0.0 – 9.7% |
| Others | 51 (14.8%) | 11.2 – 19.0% | 4 (11.1%) | 3.1 – 26.1% |
| No response | 8 |  | 0 |  |
| **Vaccine type** |  |  |  |  |
| Bivalent (BA.1 or BA.5) | 178 (50.4%) | 45.1 – 55.8% | 22 (61.1%) | 43.5 – 76.9% |
| Ancestral monovalent | 156 (44.2%) | 38.9 – 49.5% | 14 (38.9%) | 23.1 – 56.5% |
| Unvaccinated | 19 (5.4%) | 3.3 – 8.3% | 0 (0.0%) | 0.0 – 9.7% |
| **Prior infection history** |  |  |  |  |
| No prior-Infection | 350 (99.2%) | 97.5 – 99.8% | 0 (0.0%) | 0.0 – 9.7% |
| Prior-infection | 3 (0.8%) | 0.2 – 2.5% | 36 (100%) | 90.3 – 100.0% |
| **Newly diagnosis** |  |  |  |  |
| Newly diagnosed | 180 (51%) | 45.6 – 56.3% | 2 (5.6%) | 0.7 – 18.7% |
| Non newly diagnosed | 173 (49%) | 43.7 – 54.4% | 34 (94.4%) | 81.3 – 99.3% |
| **Newly anti-N positive ($\geq 1$ COI)** |  |  |  |  |
| Already anti-N positive | 2 (0.6%) | 0.1 – 2.0% | 35 (97.2%) | 85.5 – 99.9% |
| Newly anti-N positive | 341 (96.6%) | 94.1 – 98.2% | 1 (2.8%) | 0.1 – 14.5% |
| Anti-N negative | 10 (2.8%) | 1.4 – 5.2% | 0 (0.0%) | 0.0 – 9.7% |
| **Anti-N fold increase** |  |  |  |  |
| $\geq 4$ fold increase | 350 (99.2%) | 97.5 – 99.8% | 35 (97.2%) | 85.5 – 99.9% |
| $< 4$ fold increase | 3 (0.8%) | 0.2 – 2.5% | 1 (2.8%) | 0.1 – 14.5% |
| **Subjective symptoms** |  |  |  |  |
| Symptomatic | 167 (47.3%) | 42.0 – 52.7% | 2 (5.6%) | 0.7 – 18.7% |
| Asymptomatic | 12 (3.4%) | 1.8 – 5.9% | 0 (0.0%) | 0.0 – 9.7% |
| Unaware of any symptoms<br>and infection | 166 (47%) | 41.7 – 52.4% | 23 (63.9%) | 46.2 – 79.2% |
| Unknown/No response | 8 (2.3%) | 1.0 – 4.4% | 11 (30.6%) | 16.3 – 48.1% |

### Estimating the infection prevention efficacy, including re-infections, based on anti-S and anti-N antibody levels

The absolute risk reduction in newly infected individuals with or without hybrid immunity during the study period, which was the Omicron BA.5 endemic period, by combined serum anti-S and anti-N antibody levels at the initial test was estimated using a generalized additive model with inverse probability weighting (**Fig. 3A, B**). The conditional effect of the anti-N antibody levels indicated that the risk of infection decreased logarithmically with increasing anti-N antibody levels (**Fig. 3A**). For reference value, individuals with more than 10·1 COI anti-N antibody levels, which is close to the median (12.2 COI) of those of hybrid immunity holders at the initial test, were estimated to have an 80% relative risk reduction of new infections compared with a control group with no vaccination or prior infection history. As shown above, anti-N antibody levels peaked 1-2 months post-infection and declined over time (**Fig. 2D**). The estimated median duration for which anti-N antibody levels remained over 10·1 COI was 193 (CredI, 179–211) days after primary infection and 680 (CredI, 355–>1000) days after re-infection (**Fig. 2D**). While higher anti-S antibody titers were associated with a decreased risk of infection, the impact of this relationship was modest (**Fig. 3A**). Even when the anti-S antibody titer reached its highest level of 523,000 BAU/mL, the estimated reduction in relative risk was 75%. Evaluation of the combined impact of anti-N and anti-S antibody levels revealed a modest decrease in the absolute risk of infection attributable to anti-S antibody titers, whereas a reduction in the absolute risk of infection was significantly associated with an increase in anti-N antibody levels (**Fig. 3B**).

**Fig. 3.**
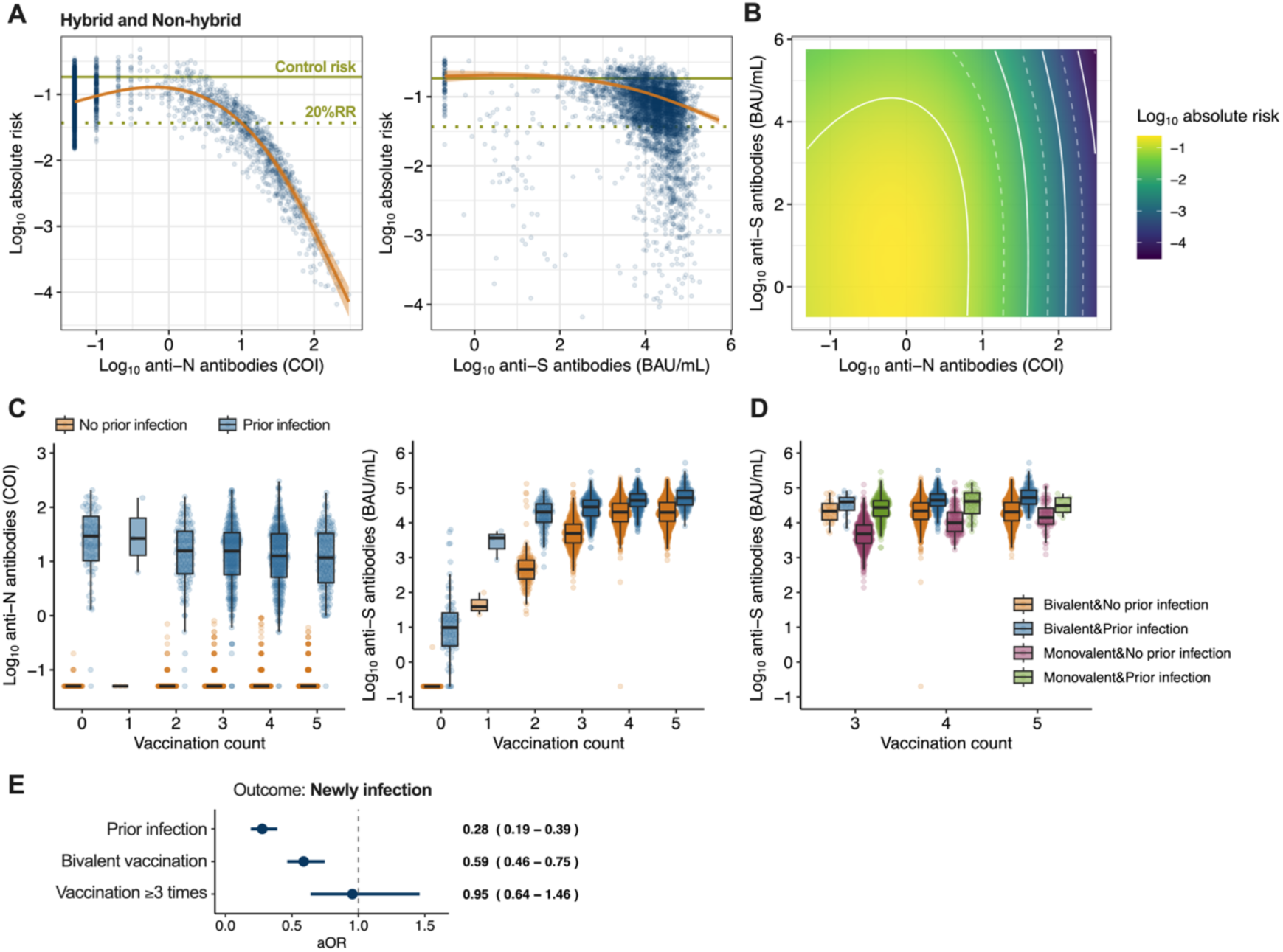
Estimation of the effect of baseline anti-N and anti-S antibody levels on infection risk during the study period. (A) The conditional effects of anti-N antibody levels (left) and anti-S antibody titers (right) during the observation period on the absolute risk of infection. Each predicted data point (blue), along with the median absolute risk and 95% credibility intervals (ribbon), are shown. The overall risk of a control group with no vaccination or prior infection history in this cohort (0·184) (green solid line) and the 80% reduction of relative risk (i.e., 20% relative risk [20%RR]) (green dotted line) are shown. (B) The combined effect of anti-N and anti-S antibody levels during the observation period on the absolute risk of infection. The logarithmic absolute risk of infection is indicated by the color bar. The white dotted and solid lines show the log_10_ absolute risk decrease for every 0·5 and 1·0, respectively. (C) Baseline anti-N antibody levels (left) and anti-S antibody titers (right) by the number of vaccine doses and infection history. (D) Baseline anti-S antibody titers by the number of vaccine doses, Omicron-adapted bivalent vaccination, and infection history. (E) Logistic regression involving factors contributing to reduction of primary and re-infection. The adjusted odds ratio (aOR) and the 95% confidence intervals are shown.

Each antibody level at the time of the initial test was assessed to determine the impact of vaccine dosage and previous infection history before the study period. Anti-N antibody levels at the time of the initial test in patients with a prior infection history were not affected by variations in the vaccination dosage (**Fig. 3C**). In contrast, the anti-S antibody titers increased as the number of vaccinations increased, and individuals who were previously infected before the study period tended to have greater anti-S antibody titers than those vaccinated, regardless of the number of vaccinations (**Fig. 3C, D**). Similarly, a higher association of prior infection, rather than Omicron-adapted bivalent vaccination or booster vaccinations, with a reduction in the risk of primary and re-infection was observed (**Fig. 3E**). These results indicate that the reduced risk of infection at high anti-S antibody titers was associated with infection-induced immunity and correlated more strongly with anti-N antibody levels than with vaccine-induced anti-ancestral S antibody titers. Finally, we estimated the absolute risk reduction for new infections, including only those with hybrid immunity, during the study period (**Fig. 4A, B**). The conditional effect of anti-N antibody levels indicated that the risk of re-infection decreased on a logarithmic scale with an increase in anti-N antibody levels, as shown in Fig. 2A (**Fig. 4A**). However, the conditional effect of anti-S antibody titers or the combined effect of anti-N antibody levels did not show a link between high anti-S antibody titers and a reduced risk of re-infection (**Fig. 4A, B**). Taken together, given that anti-N antibody levels reflect the level of the immune response induced after viral infection,^24^ it suggests that the level of serum anti-N antibodies determined using this method is a more reliable immunological correlate than the level of anti-S antibodies in determining the effectiveness of preventing SARS-CoV-2 re-infection in individuals with hybrid immunity.

**Fig. 4.**
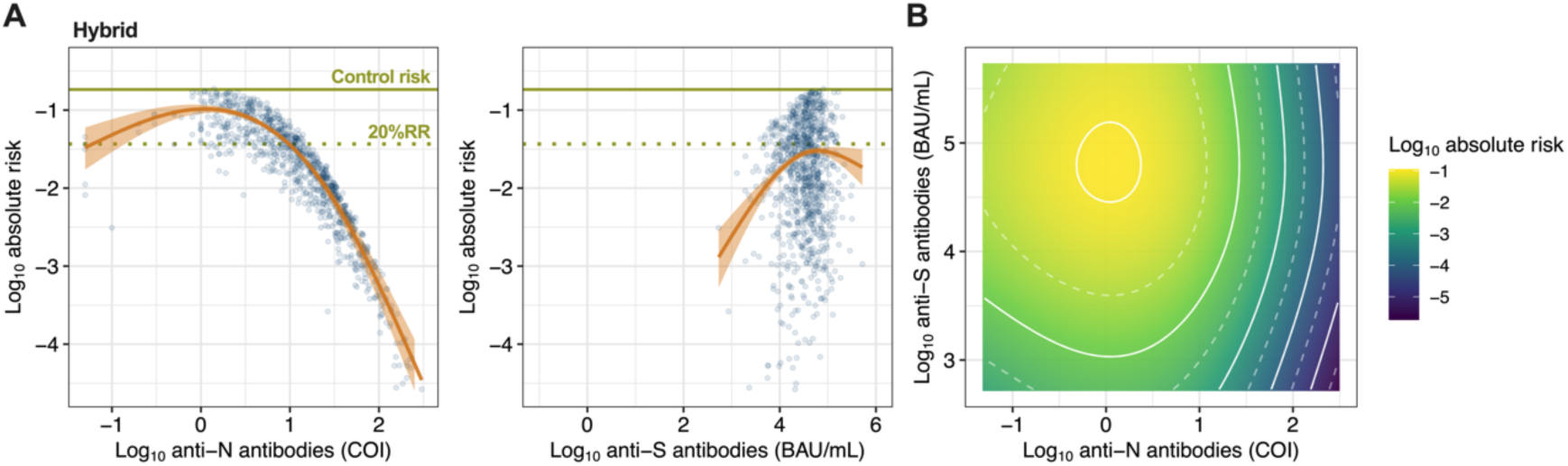
Estimation of the effect of baseline anti-N and anti-S antibody levels on re-infection risk during the study period. (A) The conditional effects of anti-N antibody levels (left) and anti-S antibody titers (right) during the observation period on the absolute risk of re-infection in individuals with hybrid immunity at baseline. The overall risk of a control group with no vaccination or prior infection history in this cohort (0·184) (green solid line) and the 80% reduction of relative risk (i.e., 20% relative risk [20%RR]) (green dotted line) are shown. (B) The combined effect of anti-N and anti-S antibody levels during the observation period on the absolute risk of re-infection.

## Discussion

This study evaluated the protective effect of serum anti-N and anti-S antibody levels against SARS-CoV-2 infection during the Omicron BA.5 epidemic period and showed that reducing re-infection risk was correlated with anti-N antibody levels, induced by previous infection, on a logarithmic scale in hybrid immunity holders with a history of infections and vaccination. In contrast, anti-S antibody titers against the ancestral spike antigen induced by both vaccines and infections showed relatively low protective effects against infection during the BA.5 epidemic period. These results suggest that the risk of re-infection can be assessed based on anti-N antibody titers in individuals with hybrid immunity. Moreover, this finding implies that infection-induced immunity in individuals with hybrid immunity includes a distinct and potent immunity beyond the presence of serum anti-S antibodies, which correlates with serum anti-N antibody levels. This would indicate the possibility for the development of next-generation COVID-19 vaccines that induce more effective immunity.

Anti-N antibodies recognize the nucleocapsid proteins inside virions or infected cells and do not directly neutralize infectious virus particles, suggesting that anti-N antibody levels are non-mechanical immunological correlates of protection (nCoPs), which are associated with some protective immunity against infection.^25^ The potential of anti-N antibody levels as immunological markers against re-infection has also been reported in children aged 4 to 15 years, consistent with the findings of this study.^26^ Immunological markers induced by infection, such as nasal mucosal S-specific secretory IgA antibodies,^17^ which are also associated with a reduction in the duration of virus shedding after infection^18^ and blood circulating N-specific CD4^+^ and CD8^+^ T cells induced by infection, but not spike-based current vaccination, associated with the prevention of infection, have been reported to negatively correlate with peak upper respiratory tract viral loads.^27^ Importantly, these immunological markers represent humoral and cellular immune responses to viral infections in the upper respiratory tract. Serum anti-N antibody responses are similarly considered to be indicative of an immune response to viral infections in the upper respiratory tract. Previously, we reported a positive correlation between the serum anti-N antibody response and upper respiratory viral load at the onset of breakthrough infections,^24^ and revealed that the upper respiratory viral load determines the magnitude of antiviral humoral immune responses after infection. Taken together, these previous findings indicate that serum anti-N antibody levels serve as a quantitative proxy for the magnitude of antiviral humoral immune responses induced after infection.

For individuals who experienced a single infection, the time for anti-N antibody levels to become negative (below 1·0 COI) was estimated to be approximately 621 days (**Fig. 2D**). These anti-N antibody waning dynamics are consistent with a recent longitudinal sampling cohort study using the same method of anti-N antibody measurement as in this study, which showed that many participants maintained positive anti-N antibody levels for more than 500 days.^22,28^ In addition, a previous report found that anti-N antibody levels were higher in re-infections than in primary infection,^23^ which is consistent with the findings of this study. Notably, the anti-N antibody levels induced by the primary infection were relatively low and below an 80% risk reduction level for re-infection at 193 days after the primary infection (**Fig. 2D**). This suggests that a single infection does not induce robust long-term immunity for re-infection prevention. This is consistent with systematic reviews indicating that the efficacy of infection prevention in previously infected individuals against re-infection with Omicron declines to approximately 50% within 20 weeks.^16^ Nasal anti-S IgA has been reported to persist for approximately nine months, and strong correlations were observed between nasal anti-S IgA and nasal anti-N IgA titers at six or 12 months after infection, but not with plasma anti-N IgA titers.^29^ Currently, the long-term relationship between serum anti-N antibodies and mucosal-specific secretory IgA or T cell immunity is unclear, and further research is needed to identify the long-term immune responses associated with anti-N antibody levels.

Several studies conducted during the pre-Omicron epidemic period following vaccine introduction have reported associations between elevated anti-S antibody or neutralizing antibody titers and strong protection against SARS-CoV-2 infection.^5–7^ Feng et al. demonstrated 80% protection against SARS-CoV-2 infection when the anti-S antibody level was 247 BAU/ml in a ChAdOx1 study cohort. During the Omicron BA.1/BA.2 epidemic period, reductions in infection risk were reported for anti-S antibody titers above 2000 BAU/mL, 800 BAU/mL, and 4810–11233 BAU/mL for symptomatic infections^9–11^ Additionally, during the Omicron BA.4/5 epidemic period, reductions in infection risk were reported for anti-S antibody titers above 380–1560 BAU/mL, but the anti-S antibody titer thresholds varied according to previous infection histories, and the association between the anti-S antibody titers and the infection risk was not observed in individuals with previous Omicron BA.2 infection.^12^ Our results did not estimate an 80% relative risk reduction against infection, even at anti-S antibody titers above 100,000 BAU/mL during the Omicron BA.5 epidemic period. This might reflect the increasing difficulty in estimating infection prevention efficacy using ancestral strain S antibody levels because of the antigenic discrepancy of Omicron sublineages that have acquired greater immune evasion capabilities and confounding factors of local upper respiratory immune responses due to prior infection.

In this study, we found that, during the Omicron BA.5 epidemic period, the reduction in re-infection risk was significantly correlated with higher anti-N antibody levels induced by prior infection. Conversely, anti-S antibody titers induced by both vaccines and infections were less strongly correlated with protection. These findings suggest that immunity correlated with anti-N antibody levels—such as mucosal antibodies, T-cell responses, and other unknown factors—may be good target immunity induced by the next-generation COVID-19 vaccines aiming the control the future COVID-19 epidemic.

### Limitations

The observation period of this study was limited to approximately two months; therefore, a longer observation period could provide a better estimation of the long-term protective effects and the precise duration of immunity for infection protection. In this study, anti-S antibody titers against Omicron spikes, which are expected to contribute to the prevention of Omicron infection, were not measured, and the correlation between anti-N antibody levels and anti-Omicron S antibody titers remains unknown. We did not evaluate the effect of antibody titers on the prevention of severe disease since the questionnaire did not include the severity in this survey, although the prevention of severe disease through vaccination has been confirmed even during the Omicron BA.1/2 and BA.4/5 epidemic periods. While the participants with a prior infection history included pre-Omicron-infected individuals, most were Omicron-infected cases. The quality of immunity induced by infection may vary with the variant, but it has not been determined which variant infection induced immunity in each prior-infected individual. We have not examined the correlation between serum anti-N antibody levels and infection prevention efficacy for variant infections outside the BA.5 epidemic period. Further investigation is needed to determine if the correlation between anti-N antibody titers and infection prevention can be applied to Omicron sublineages post-BA.5.

## Methods

### Survey design, participants and ad hoc study design

For the national COVID-19 seroepidemiological survey, residents of Miyagi, Tokyo, Aichi, Osaka, and Fukuoka prefectures were randomly selected using the Basic Resident Register via multistage sampling as described previously.^30^ For each prefecture, at least one municipality from each of the following three municipality types was chosen for the surveys: small (<100 000 population), medium (≥100 000 population), and large (ordinance-designated city/special ward). The samples were divided according to the relative population sizes of the municipalities, and residents were randomly sampled from each municipality. We planned to enroll 15,000 individuals from five prefectures (3,000 individuals per prefecture). Assuming a response rate of 20%, we randomly sampled 75,000 individuals aged 20 years or older and invited them via mail. Only one participant from each household participated in the study. No financial incentives are provided for the participants except for travel subsidy to the study sites and feedback of the serologic test results. Individuals who agreed to participate in the study answered a self-administered questionnaire, visited the designated site where they provided written consent, and had their blood drawn. Briefly, the questionnaire included the demographic information (age, biological sex, occupation, municipality, etc.), comorbidities, COVID-19 vaccination status, and history of SARS-CoV-2 infection. The survey was conducted as a public health investigation under the Act on the Prevention of Infectious Diseases and Medical Care for Patients with Infectious Diseases (Infectious Diseases Control Law) and planned by the Ministry of Health, Labor, and Welfare of Japan (MHLW) and the National Institute of Infectious Disease (NIID). The survey assessed the prevalence of anti-N and anti-S antibodies. The MHLW randomly selected the potential study participants and mailed invitations to them to participate in the survey, which was carried out as a public health investigation under the Infectious Diseases Control Law. Descriptive results of the national COVID-19 seroepidemiological survey for the entire cohort have been published in Japanese on the MHLW/NIID websites. ^31^

The ad hoc study as a research activity evaluated the protective efficacy of anti-N antibodies against reinfection, based on the survey data. Consent for this ad hoc study was obtained as part of the survey consent process, which included agreement to use data for further research.

### Ethical approval

All the samples, protocols, and procedures described herein were approved by the Medical Research Ethics Committee of the NIID and involved human participants, based on the principles of the Declaration of Helsinki (approval numbers 1457, 1472 and 1730).

### Electrochemiluminescence immunoassay

Serum samples were heat-inactivated at 56 °C for 30 min before use. Antibody titers for the ancestral spike (S) receptor-binding domain and nucleocapsid (N) were measured using Elecsys Anti-SARS-CoV-2 S (Roche, Basel, Switzerland) and Elecsys Anti-SARS-CoV-2 (Roche) kits, respectively, according to the manufacturer’s instructions. A cutoff index (COI) of 1·0 and a cutoff value of 0·8 BAU/ml, as determined by the manufacturer, were used to determine the presence/absence of anti-N antibody levels and anti-S antibody titers, respectively. Since the COVID-19 vaccines approved in Japan are only spike-based vaccines, anti-N antibodies are induced by infection but not by vaccines, whereas anti-S antibodies are induced by both infection and vaccines. The numerical results in U/mL of the Elecsys Anti-SARS-CoV-2 S assay and the WHO BAU/mL are equivalent.

### Definition of newly infected individuals

The study included 5,627 individuals who participated in two consecutive tests. Those who were newly vaccinated during the observation period were excluded (1,044 individuals). To use anti-N antibody seroconversion to determine infection, we excluded 87 individuals with a history of infection within 30 days of the initial antibody test (87 individuals). Consequently, there were 4,496 subjects eligible for the study.

Participants who tested positive for COVID-19, who were diagnosed after the baseline initial test date, or who had an anti-N titer turn positive to 1·0 COI or higher were considered newly infected. In addition, participants who showed a four-fold or higher increase in anti-N antibody levels in the second antibody test compared to baseline were considered newly infected, including those with re-infections (**Fig. 2**).

### Definition of symptomatic individuals

Symptomatic individuals were defined as individuals with any of the following, based on a previous study:^4^ malaise, chills, joint pain, headache, runny nose, cough, sore throat, shortness of breath, gastrointestinal symptoms (vomiting, diarrhea, or stomach ache), and loss of taste or smell. Those who answered that they were infected with SARS-CoV-2 but were asymptomatic were classified as “asymptomatic.” Those who did not report a SARS-CoV-2 infection or any symptoms but met the criteria for a new infection were classified as “Unaware of any symptoms and infection.”

### Infection risk estimation

The immune correlates of the infection risk analysis were conducted based on a previous study.^6^ Log-transformed anti-S and anti-N antibody titers were analyzed using Bayesian generalized additive models (GAM) for binary data, with cubic spline smoothing applied to antibody titers to allow a nonlinear effect. No adjustments were made for multiple comparisons. Combined models were fitted for anti-S and anti-N antibody levels, controlling for baseline exposure risk, and weighted using inverse probability weights as described below.

In addition to antibody titers, we assumed that each participant’s absolute risk of new infections varied depending on region and demographics. To adjust the estimates of the absolute risk of infection due to each participant’s characteristics, we used a Poisson regression model to predict the probability of infection. The predictors were age group (20–64 years or 65 years or older), biological sex, presence of comorbidities, Omicron BA.1/BA.5 bivalent vaccination, vaccination count, prior infection history, and municipality. The inverse probability from this model was used to weigh the correlates of the risk models and eliminate the source of bias. In the model with hybrid immunity alone, we excluded the municipality from the predictors to avoid excessive weight.

The newly infected response variable was modeled as a function of smooth terms for anti-S and anti-N antibody levels using cubic splines with three basis functions (k = 3). To account for the overdispersion and potential zero inflation in the count data, we specified a zero-inflation Poisson distribution for the response.

Parameter estimation was performed using the Markov chain Monte Carlo (MCMC) approach implemented in *rstan* 2.26 (https://mc-stan.org). Four independent MCMC chains were run with 5,000 steps including a warm-up period of 1,000 steps, with subsampling every five iterations. We confirmed that all the estimated parameters showed <1·01 R-hat convergence diagnostic values and >1600 effective sampling size values, indicating that the MCMC runs were convergent. Information on the model estimates is summarized in **Tables S1 and S2**.

### Modeling the antibody response

To model the anti-N antibody response in individuals diagnosed with the first infection and re-infection with SARS-CoV-2, a Bayesian model was used based on a previous study.^32^ The measurement (*i.e.,* log_10_ anti-N) *y_i_* was modeled as *y_i_* ∼ *Normal(h f(t_i_, α, β)* exp(-*λt_i_*), *σ*), where *Normal(a, b)* indicates normal distribution with mean *a* and standard deviation *b*, *t_i_* is the time of the measurement, *f*(*t_i_, α, β*) is the cumulative gamma distribution function at time *t_i_* with shape α and inverse scale *β, λ* is the decay rate, *σ* is standard deviation of the normal distribution, and *h* is the maximum response if *λ* = 0.^32^ The posterior distribution of each parameter was sampled for each infection group. For the prior distribution of *h*, we used weakly informed priors, *Normal*(0, 5). For the prior distributions of *α, β, λ*, and *σ*, we used a Student’s t distribution with four degrees of freedom, instead of a normal distribution, to reduce the effects of outlier values.^33^

Parameter estimation was performed using an MCMC approach implemented in *rstan*. Four independent MCMC chains were run with 4,000 steps including a warm-up period of 2,000 steps, with subsampling at every five iterations. We confirmed that all estimated parameters showed <1·01 R-hat convergence diagnostic values and >1000 effective sampling size values, indicating that the MCMC runs were convergent. The estimated means are summarized in **Table S3**.

### Statistical analysis

We performed logistic regression with the high anti-S antibody titer holder as the outcome, adjusting for age group (20–64 years or 65 years or above), biological sex, prior-infection history, bivalent vaccination, and ≥3 times vaccination. The confidence intervals (CIs) of the categorical variables in the tables were calculated using two-sided Fisher’s exact test. Pearson’s correlation coefficients were used to assess correlations between continuous variables. All statistical analyses were performed using R (version 4.3), and antibody titers below the detection limit were converted to half of the detection limit. The infection risk estimation model was constructed using *brms* 2.20.

## Data Availability

Research data reported in this paper is available from the corresponding author upon request.

## Authors’ contributions

Conceptualization, SM, RK, TA, HT, HH, SA, MS, DY, TW, and TS; Methodology, SM, RK, TA, DY, and TS; Research investigation, SM, KN, RK, TA, MH, MS, DY, TW, and TS; Public health investigation, SM, RK, HT, HH, TK, AS, AA, SA, MS, DY, TW, and TS; Data curation, SM, KN, MH and TS; Computational analysis, SM and KN; Formal analysis, SM, KN, MH, and TS; Visualization, SM; Research funding acquisition, SM, RK, MS, DY, TW, and TS; Project administration, MS, and TS; Supervision, MS, YD, and TS; Writing the original draft, SM and TS; Writing – review & editing, SM, KN, RK, MS, DY, and TS. All authors agreed to submit the manuscript, read and approved the final draft, and take full responsibility for its content, including data accuracy and statistical analysis.

## Data sharing

Research data reported in this paper is available from the corresponding author upon request.

## Acknowledgments

We thank the Miyagi, Tokyo, Aichi, Osaka, and Fukuoka prefecture governments for their support in implementing the national COVID-19 seroepidemiological survey. We also thank Shoko Sakuraba and Jun Sugihara for support in implementing the survey from the Ministry of Health, Labour and Welfare, Japan. We also thank the staff members at the Survey Research Center, Mitsubishi Research Institute, SRL, Inc., and Benefit One Inc. for their administrative and technical assistance for the survey. The national COVID-19 seroepidemiological survey was funded by the MHLW as a public health investigation. The ad hoc study based on the survey data as a research activity was supported in part by a Grant-in-Aid for JSPS Scientific Research (KAKENHI) (23K27422 (to TS), 21K20768 (to SM), 23K14534 (to SM), and 21K17307 (to RK)); AMED Research Program on Emerging and Re-emerging Infectious Diseases (JP23fk0108637 (to TS), JP22fk0108509 (to TS), JP 23fk0108684 (to TS), and JP22fk0108568 (to SM)); MHLW Emerging/Reemerging Infectious Diseases and Vaccination Policy Promotion Research Project (24HA2009 (to TS), 22HA2006 (to TS), 21HA2005 (to TS) and 20HA2001 (to TS)); the JST, PRESTO (JPMJPR21RC (to DY)).

## Supplementary Materials

**Table S1.**
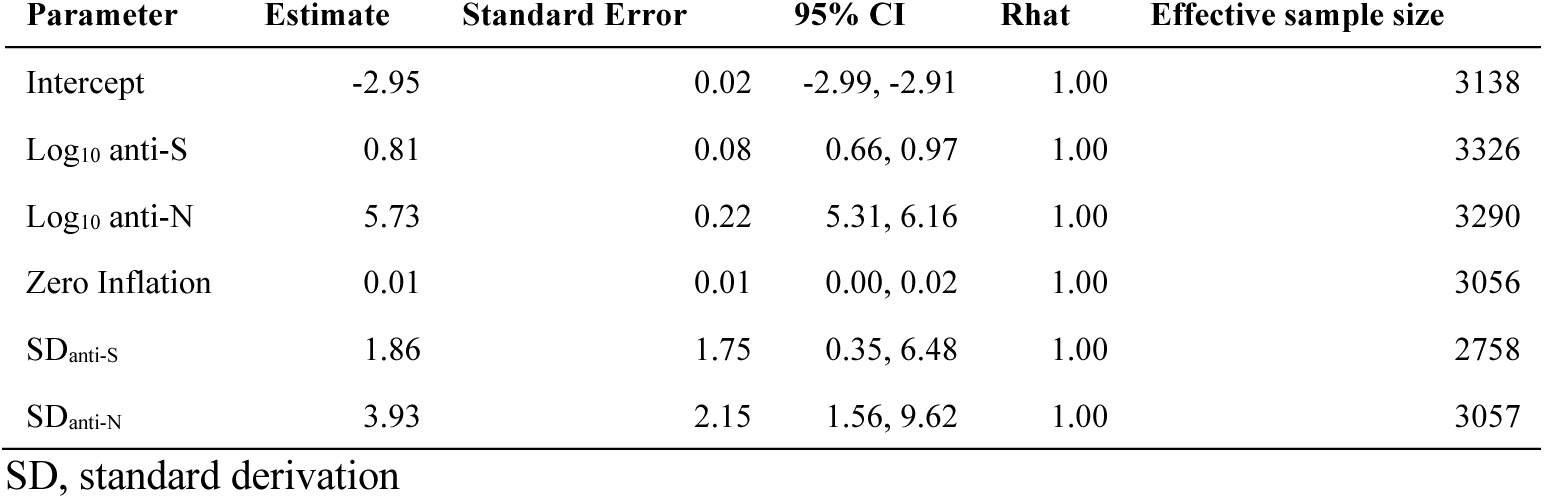
Summary of the model estimates from total infection risk estimation.

**Table S2.** Summary of the model estimates from re-infection risk estimation.

| Parameter | Estimate | Standard Error | 95% CI | Rhat | Effective sample size |
| --- | --- | --- | --- | --- | --- |
| Intercept | -4.20 | 0.07 | -4.35, -4.06 | 1.00 | 3184 |
| Log<sub>10</sub> anti-S | -2.53 | 0.31 | -3.15, -1.94 | 1.00 | 3187 |
| Log<sub>10</sub> anti-N | 3.70 | 0.20 | 3.29, 4.09 | 1.00 | 3412 |
| Zero Inflation | 0.03 | 0.02 | 0.00, 0.09 | 1.00 | 3033 |
| SD<sub>anti-S</sub> | 2.51 | 1.92 | 0.61, 7.39 | 1.00 | 3126 |
| SD<sub>anti-N</sub> | 4.47 | 2.57 | 1.76, 11.6 | 1.00 | 3088 |

SD, standard derivation

**Table S3.** Summary of the model estimates from anti-N antibody response.

| Infection history | Parameter | Estimate | Standard Error | 95% CI | Rhat | Effective sample size |
| --- | --- | --- | --- | --- | --- | --- |
| Primary infection | $h$ | 3.16 | 0.00 | 3.02, 3.36 | 1.00 | 1416 |
| Primary infection | $\alpha$ | 0.59 | 0.00 | 0.41, 0.81 | 1.00 | 1425 |
| Primary infection | $\beta$ | 0.04 | 0.00 | 0.02, 0.06 | 1.00 | 1409 |
| Primary infection | $\lambda$ | 0.00 | 0.00 | 0.00, 0.00 | 1.00 | 1469 |
| Primary infection | $\sigma$ | 0.56 | 0.00 | 0.54, 0.59 | 1.00 | 1581 |
| Re-infection | $h$ | 3.09 | 0.00 | 2.76, 3.51 | 1.00 | 1532 |
| Reinfection | $\alpha$ | 0.47 | 0.01 | 0.02, 1.57 | 1.00 | 1733 |
| Reinfection | $\beta$ | 0.51 | 0.01 | 0.05, 1.70 | 1.00 | 1557 |
| Reinfection | $\lambda$ | 0.00 | 0.00 | 0.00, 0.00 | 1.00 | 1580 |
| Reinfection | $\sigma$ | 0.52 | 0.00 | 0.39, 0.69 | 1.00 | 1612 |

